# Use of a digital adherence technology for tuberculosis treatment supervision among adolescents

**DOI:** 10.1101/2024.11.22.24316602

**Authors:** Ann M Schraufnagel, Rebecca Crowder, Peter Wambi, Suzan Nakasendwa, Alex Kityamuwesi, Devan Jaganath, Muhammad Musoke, Joyce Nannozi, Joseph Waswa, Agnes Nakate Sanyu, Maureen Lamunu, Amon Twinamasiko, Lynn Kunihira Tinka, Denis Oyuku, Diana Babirye, Christopher Berger, Ryan Thompson, Stavia Turyahabwe, David Dowdy, Achilles Katamba, Adithya Cattamanchi, Noah Kiwanuka

**Affiliations:** Division of Pulmonary and Critical Care Medicine, San Francisco General Hospital, University of California San Francisco, San Francisco, USA; Center for Tuberculosis, University of California San Francisco, San Francisco, USA; Mulago National Referral Hospital, Kampala, Uganda; Uganda Tuberculosis Implementation Research Consortium, Kampala, Uganda; Department of Epidemiology, Johns Hopkins Bloomberg School of Public Health, Baltimore, USA; National Tuberculosis and Leprosy Program, Uganda Ministry of Health, Kampala, Uganda; Clinical Epidemiology & Biostatistics Unit, Department of Medicine, Makerere University College of Health Sciences, Kampala, Uganda; Division of Pulmonary Diseases and Critical Care Medicine, University of California Irvine, Irvine, USA; Department of Epidemiology and Biostatistics, School of Public Health, Makerere University College of Health Sciences, Kampala, Uganda

## Abstract

**Background:** Adolescents (ages 10-19) affected by tuberculosis (TB) face unique challenges to completing TB treatment, resulting in increased loss to follow-up and mortality as compared to younger children and older adults. Digital adherence technologies (DATs) may be a useful tool for TB treatment monitoring. In this study we aimed to assess whether 99DOTS, a low-cost DAT, could improve treatment outcomes among adolescents with pulmonary TB (PTB).

**Methods:** We conducted an interrupted time series (ITS) analysis of adolescents initiating treatment for drug susceptible PTB between August 1, 2019 and June 30, 2021 at 30 health facilities in Uganda. ITS analysis was used to model the change in TB treatment outcomes and loss to follow-up in adolescents prior to and after the implementation of a 99DOTS-based intervention. Immediate (level) and trend (slope) changes in outcomes were assessed according to intention-to-treat (ITT) and per protocol (PP) principles.

**Findings:** 630 adolescents were included in the ITT analysis. In the post-intervention period, 78.4% of adolescents were enrolled on 99DOTS. The proportion successfully completing TB treatment was 92.4% (303/328) in the post-intervention period and 87.4% (264/302) in the pre-intervention period. In the adjusted ITT analysis, the proportion treated successfully increased (level change, PR 1.18 95% CI 1.08-1.28) and the proportion lost to follow-up decreased (level change, PR 0.93, 95% CI 0.88-0.98) in the immediate post-intervention period. Both proportions remained similar throughout the post-intervention period (p-value for slope change >0.05).

**Interpretation:** There was a high uptake of 99DOTS among adolescents with TB, and use of 99DOTS was associated with improved treatment outcomes. DATs should be further explored as a promising adolescent-specific tool for improving treatment outcomes among adolescents with TB.

## INTRODUCTION

Adolescence, a critical period during the life course, is a particularly consequential time to be ill with tuberculosis (TB). Following a relatively lower incidence during mid-childhood (ages 5-9), there is a large increase in TB incidence affecting adolescents (ages 10-19), with an estimated 850,000 adolescents affected worldwide each year.^1^ Adolescence is an important period of biological growth and transition in social roles, and having TB disease can have significant consequences. Insufficient nutrition, as is often seen in TB disease,^2,3^ can impair endocrine factors critical for normal adolescent growth and development.^4^ The effects of isolation, stigma, and discrimination during adolescence can have long-lasting social and emotional consequences.^1,5^ Missed school due to illness, directly observed therapy (DOT) visits^6^, and school- or government-induced periods of isolation^7,8^ can further impact long-term career prospects and prosperity.^8^

Despite these compelling reasons to cure adolescents with TB, adolescents experience increased rates of loss to follow-up and mortality as compared to adults and children ages 1-9. ^9–12^ They face unique barriers to completing TB treatment given an increased sensitivity to stigma, feelings of invulnerability to TB, and comorbid depression and substance use disorders.^7,13^ Further issues with medication adherence can arise when adolescents have complicated caregiver support, for example, when access to DOT clinics are geographically limited by divorced parents who live in different communities.^7^ A lack of adolescent-friendly TB clinic services and care, such as no age-appropriate TB education and adherence counseling, adolescent-friendly clinic spaces separated from older adults, and lack of monitoring approaches that does not conflict with school or work hours, further hinder treatment adherence.^13,14^

Digital adherence technologies (DATs) may be a particularly helpful TB treatment monitoring tool for adolescents given their high technology use and mobile phone access^15^, but there are limited studies examining these technologies in adolescents. 99DOTS is a low-cost DAT whereby people with TB self-report medication dosing by calling a toll-free phone number hidden underneath pills in blister packs^16,17^ and has been trialed in countries including India,^17,18^ Uganda,^16^ and Kenya.^19^ In a previous study of 99DOTS among adults with TB in Uganda, we showed that provision of low-cost phones and engaging community health workers’ support markedly increased enrollment on 99DOTS, but that 99DOTS-based treatment supervision did not improve treatment outcomes.^20^

In line with World Health Organization’s (WHO) call for adolescent-centered services,^21^ we aimed to assess whether 99DOTS-based treatment supervision could improve treatment outcomes among adolescents with drug-susceptible pulmonary TB (PTB).

## METHODS

### Study design and population

We conducted an interrupted time series (ITS) study to evaluate the effect of a 99DOTS-based treatment support intervention for adolescents with PTB at 30 health facilities in Uganda. These included 5 regional hospitals, 10 district hospitals, 7 Level IV (outpatient and inpatient) health centers, and 8 Level III (outpatient and maternity services) health facilities. Sites were selected based on volume of people of all ages treated for PTB (diagnosed >10 people with PTB per month in 2017), low historic treatment success rate (PTB treatment success rate of <80% in 2017), and geography (all sites within 225 kilometers of Kampala city). Results among adults (ages 18 and older) in this population were published previously.^20^

Here, we included all adolescents (defined as people ages 10-19) initiating treatment for drug susceptible PTB between August 1, 2019, and June 30, 2021. We excluded adolescents with TB who were transferred between facilities during the treatment period, as they would not have had the opportunity to fully benefit from the intervention.

### 99DOTS-based Intervention

The 99DOTS platform^16,18,22^ included automated dosing reminders via SMS and dosing confirmation via toll-free calls. To increase uptake of 99DOTS and facilitate treatment supervision, we provided low-cost phones ($8 USD) to adolescents with TB who lacked regular access to a phone, task-shifted adherence monitoring and follow-up from clinicians to community health workers (CHWs), and provided automated task lists to facilitate CHW follow-up of dosing history.

### Intervention periods

From August 2019 to April 2020 (the *pre-intervention period*), most adolescents less than 18 years old received TB treatment supervision in the same manner as before the trial. This previously described^23^ community-based DOT model varied substantially between facilities, but typically relied on a family member or other untrained treatment supporter to directly observe patients taking medications. During the pre-intervention period, the 99DOTS platform was available to older adolescents at 18 of the 30 health facilities. From May to June of 2020 (*buffer period)*, study staff conducted training on the 99DOTS-based intervention. From July 2020 to June 2021 (*post-intervention period*), the 99DOTS-based intervention was available for TB treatment support and supervision for all adolescents at all 30 health facilities (See **Appendix Figure 1**).

### Procedures

Data on demographic and clinical characteristics, as well as TB treatment initiation and outcomes, were collected from routine TB treatment registers at each health facility. Facility staff took and uploaded photos of the register to a secure, password protected server. Research staff then extracted data from these photos into a secure database using Research Electronic Data Capture (REDCap) software.^24^ Enrollment onto 99DOTS was confirmed by merging this REDCap database with a list of enrolled participants from the 99DOTS server.

### Outcomes

The primary outcome was the proportion treated successfully, defined as having a treatment outcome of cured or completed entered into the Unit Treatment Register or District Treatment Register. Secondary outcomes included the proportion lost to follow-up and the proportion of people with TB enrolled on 99DOTS during the post-intervention period. The former was defined as having a treatment outcome of lost to follow-up entered into the Unit Treatment register or District Treatment Register, and the latter as the percentage of participants enrolled on 99DOTS within twenty-eight days of initiating TB treatment during the post-intervention period.

### Statistical analysis

ITS analysis was used to model the change in TB treatment outcomes and loss to follow-up in adolescents at 30 health facilities prior to and after the implementation of the enhanced 99DOTS-based intervention. We assessed the change in the level of outcomes immediately after the buffer period of introducing the intervention (level) and trend (slope) changes in outcomes throughout the post-intervention period.

The primary and secondary outcomes were analyzed according to both intention-to-treat (ITT) and per protocol (PP) principles. The ITT analysis included all eligible participants ages 10-19. The PP analysis excluded those who did not enroll on 99DOTS, the intended method of TB treatment supervision, during the post-intervention period.

We inspected trajectory plots of observed and predicted outcomes to assess the overall post intervention linearity. Single group ITS models were fit using the mean proportion with each outcome of interest by a one-month period (see **Supplemental Methods** in **Appendix** for equation) using the Stata command *itsa*.^25^ Autocorrelation was assessed using the Cumby-Huizinga test. Proportion ratios (PR) from the ITS model characterized the pre- and post-intervention slopes, as well as the changes in level and slope difference. For adjusted analyses, we built a multivariable model using logical forward selection and included HIV status and diagnosis type (clinical versus bacteriological confirmation) as covariates. Stata version 17 was used for all analyses.

## RESULTS

### Study population

From August 1, 2019, to June 30, 2021, 943 adolescents initiated treatment for PTB at the 30 health facilities (Figure 1). The ITT analysis included 630 people (302 in the pre-intervention and 328 in the post-intervention periods) after excluding adolescents who were ineligible or treated during the buffer period. The PP population included 559 people after excluding 71 adolescents who were not enrolled on 99DOTS during the post-intervention period.

**Figure 1.**
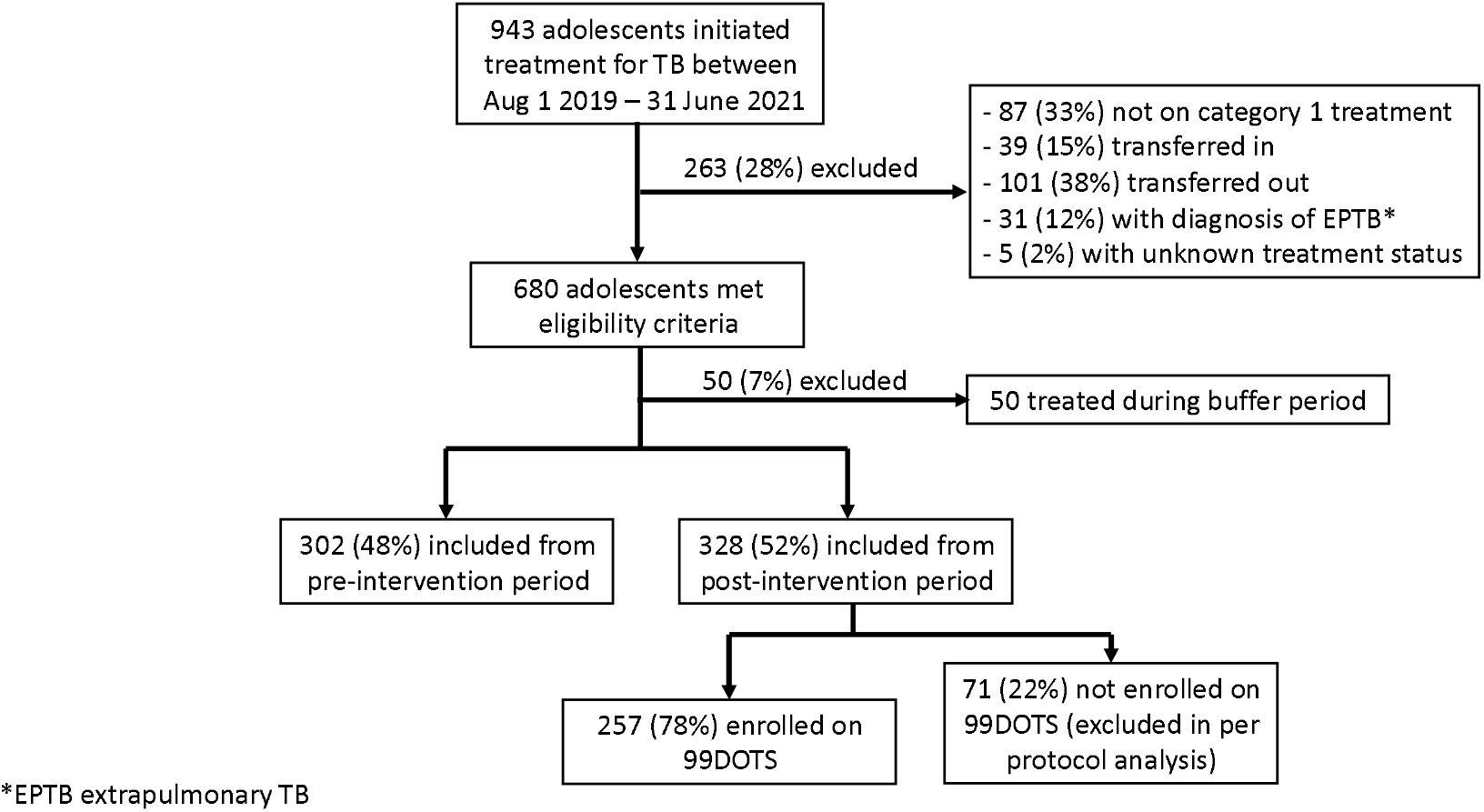
Target and analysis population.

There were no differences between the pre- and post-intervention period ITT population in median age or proportions living with HIV, on ART, or previously diagnosed with TB. The proportions female, with bacteriologically confirmed TB, and treated at a health center (as compared to a hospital) were all higher in the post-intervention period (**Table 1**). These same differences held true for the PP population; additionally, adolescents in the post-intervention PP population were older.

**Table 1.**
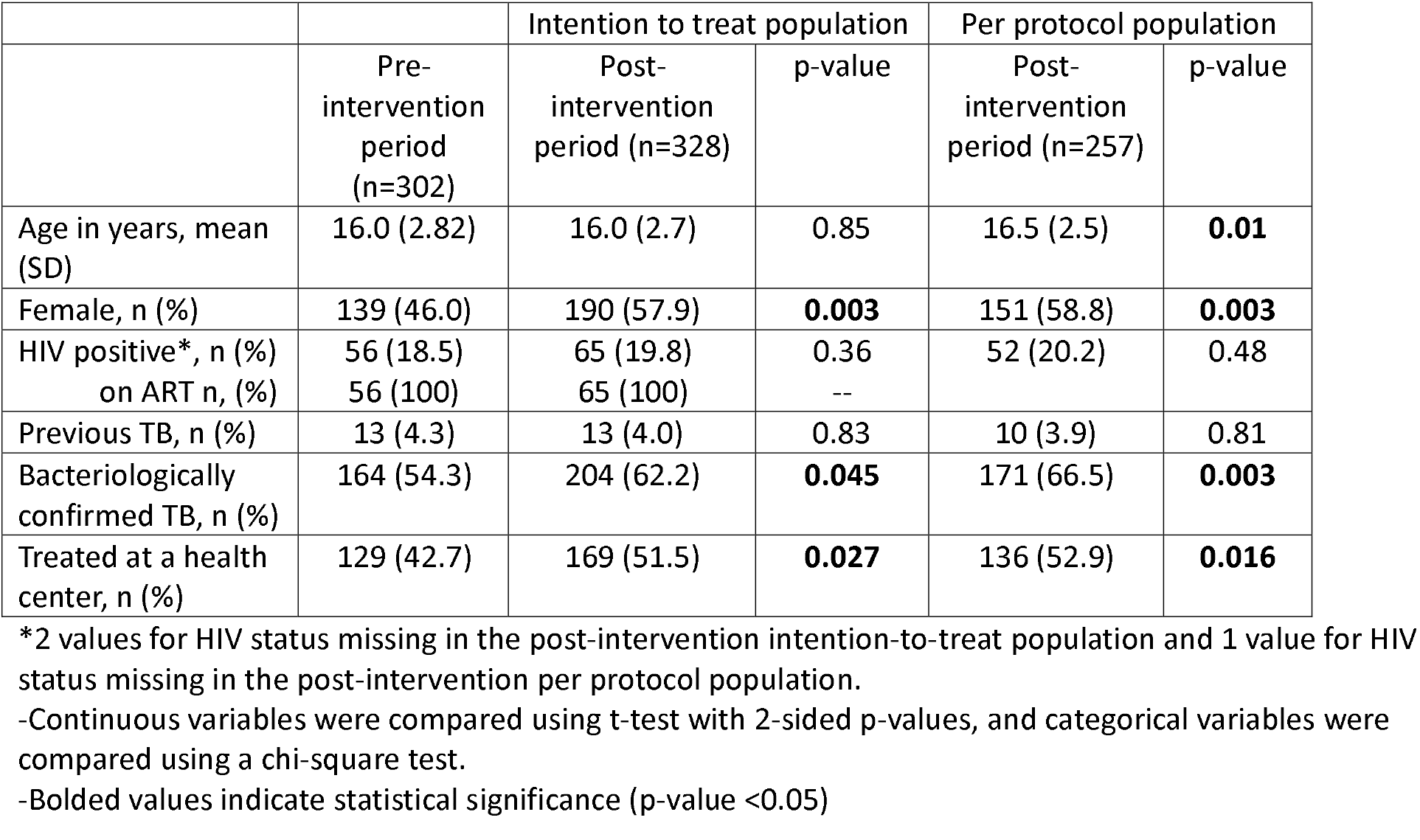
Participant baseline characteristics by study population and period.

During the pre-intervention period, 50 (19.8%) adolescents were enrolled on 99DOTS, all of whom were 18 or 19 years old (see **Appendix Figure 1**). During the post-intervention period, 257 (78.4%) adolescents were enrolled on 99DOTS within 28 days of initiating treatment for TB (**Table 2**). There was no difference in 99DOTS enrollment by sex, HIV status, previous TB, or treatment location (hospital vs health center). Adolescents with bacteriologically confirmed TB were more likely to be enrolled on 99DOTS than those with a clinical diagnosis. There were marked differences in 99DOTS enrollment by age: 43% of the youngest (10 years) as compared to 95% of the oldest (19 years) adolescents were enrolled.

**Table 2.** Characteristics of participants receiving 99DOTS during the post-intervention period.

| Characteristic | Received 99DOTS<br>N=257 (78.4%) | p-value |
| --- | --- | --- |
| Age n, (%) |  | **<0.001** |
| 10 | 6 (42.9) |  |
| 11 | 10 (55.6) |  |
| 12 | 11 (64.7) |  |
| 13 | 8 (44.4) |  |
| 14 | 17 (68.0) |  |
| 15 | 18 (72.0) |  |
| 16 | 28 (84.9) |  |
| 17 | 40 (80.0) |  |
| 18 | 59 (90.8) |  |
| 19 | 60 (95.2) |  |
| Female n, (%) | 151 (79.5) | 0.56 |
| Male n (%) | 106 (76.8) |  |
| HIV* |  | 0.59 |
| Positive n, (%) | 52 (80.0) |  |
| Negative n, (%) | 204 (78.2) |  |
| Treatment |  | 0.89 |
| New treatment n, (%) | 247 (78.4) |  |
| Retreatment n, (%) | 10 (76.9) |  |
| Bacteriologically confirmed<br>n, (%) | 171 (83.8) | **0.002** |
| Clinically diagnosed<br>n, (%) | 86 (69.4) |  |
| Received treatment at: |  | 0.34 |
| Hospital | 121 (76.1) |  |
| Health Center | 136 (80.5) |  |

\*2 missing values for HIV status
-Bolded values indicate statistical significance (p-value <0.05)

There were no missing outcome data, but two people with TB treated in the ITT population did not have a recorded HIV status as they did not undergo HIV testing.

### Primary outcomes

In the post-intervention period, 303 (92.4%) adolescents successfully completed TB treatment, as compared to 264 (87.4%) adolescents in the pre-intervention period. In the ITT analysis, after adjusting for HIV status and diagnosis, the proportion treated successfully increased in the immediate post-intervention period (level change, PR 1.18 95% CI 1.08-1.28) (**Figure 2, Table 3**). There was no change in slope following the intervention (PR 1.01, 95% CI 0.99-1.02), and the proportion treated successfully remained similar throughout the post-intervention period (PR 0.99, 95% CI 0.99-1.00). After adjusting for HIV status and diagnosis, the proportion lost to follow up decreased in the immediate post-intervention period (level change, PR 0.93, 95% CI 0.88-0.98), and the proportion lost to follow up remained similar throughout the post-intervention period (PR 1.00, 95% CI 0.99-1.01). Results were similar in the PP analysis, and with adjusted and unadjusted models (See **Supplemental Table 1**).

**Table 3.** Interrupted time series regression for all adolescents using an adjusted model.

|  | Change in level<br>(PR, 95% CI) | Change in slope<br>(PR, 95% CI) | Pre-intervention<br>slope (PR, 95% CI) | Post-intervention<br>slope (PR, 95% CI) |
| --- | --- | --- | --- | --- |
| Treated successfully |  |  |  |  |
| ITT | 1.18 (1.08-1.28)* | 1.01 (0.99-1.02) | 0.98 (0.97-1.00)* | 0.99 (0.99-1.00) |
| PP | 1.18 (1.09-1.28)* | 1.01 (1.00-1.03) | 0.98 (0.97-1.00)* | 1.00 (0.99-1.00) |
| Loss to follow-up |  |  |  |  |
| ITT | 0.93 (0.88-0.98)* | 1.00 (0.99-1.01) | 1.01 (1.00-1.02) | 1.01 (1.00-1.01) |
| PP | 0.93 (0.89-0.97)* | 1.00 (0.99-1.01) | 1.01 (1.00-1.01) | 1.00 (1.00-1.01) |

PR: proportion ratio; CI: confidence interval; ITT: intention-to-treat; PP: per protocol analysis Statistically significant results noted with \* (<0.05)
Model adjusted for HIV status and diagnosis type (clinical versus bacteriological confirmation).

**Figure 2.**
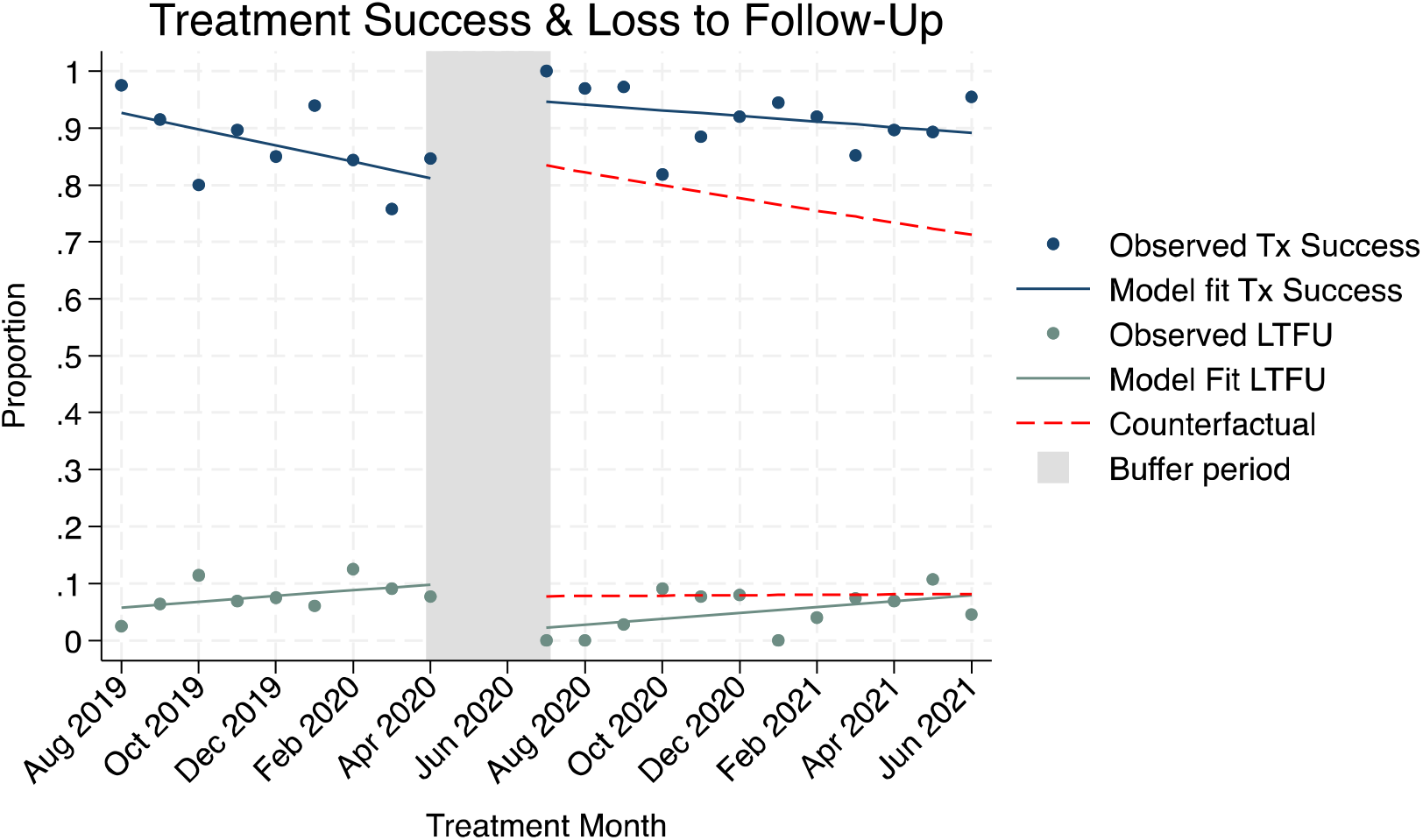
Treatment success and loss to follow-up for all adolescents

### Subgroup analyses

A subgroup analysis was performed among older adolescents (ages 15-19) given their overall high uptake of the intervention (95% enrolled on 99DOTS). In an unadjusted model, the proportion of older adolescents treated successfully increased in the immediate post-intervention period (level change, PR 1.26, 95% CI 1.11-1.43) (See Supplemental Figure 2, Table 2). There was no change in slope following the intervention (PR 1.02, 95% CI 1.00-1.04), and the proportion treated successfully remained similar throughout the post-intervention period (PR 0.99, 95% CI 0.99-1.00). The proportion lost to follow up decreased in the immediate post-intervention period (level change, PR 0.93, 95% CI 0.88-0.98), and the proportion lost to follow up remained similar throughout the post-intervention period (PR 1.00, 95% CI 0.99-1.01). Results were similar in the PP analysis (See **Supplemental Table 2**).

## DISCUSSION

In the first study to evaluate DATs for adolescents with TB, we found high uptake particularly among older adolescents and improved treatment outcomes in this important but often neglected sub-population. This is also among the first studies to demonstrate an improvement in treatment outcomes with a low-cost DAT, highlighting that DATs may be a particularly relevant alternative to DOT for adolescents. Low-cost DATs may fall within the WHO’s charge to provide adolescent-specific therapies for TB, and National TB Control Programs could consider offering them to adolescents as an alternative to DOT for treatment monitoring.

Previous studies evaluating the effect of DATs on medication adherence and treatment outcomes among adults with TB have shown mixed results: A 2022 systematic review found 4 randomized controlled trials (RCTs) with significant effects and 6 RCTs with no significant effects.^26^ Adolescents have been shown to have high technology use and mobile phone access,^15^ which may explain their high uptake of and improved outcomes with 99DOTS. 78% of all adolescents and 95% of older adolescents used 99DOTS in the post-intervention period. This compares to 86-87% of adults enrolled on 99DOTS in this population^20^ as well as previously published studies of 99DOTS in adults, where approximately one third to one half of adults used the technology.^16,17^

Further studies are needed to better understand the differential uptake of DATs between younger and older adolescents as was seen in this study. It is plausible that younger adolescents have lower access to mobile phones and higher dependence on caregivers for their medical treatment decisions. However, if messaging is geared at older adolescents or adults, this could be an additional factor limiting younger adolescents’ uptake of the technology. Additional qualitative work in this area, as well as stratifying adolescence into “young” (10-14 years), “middle” (15-19 years) and “late” adolescence (20-24 years) as per experts recommendations^27^ could help better understand unique barriers to treatment and to develop more nuanced age-appropriate interventions.

There were some limitations to this study. In ITS analyses, results can be impacted by time-varying confounders that do not remain consistent across the pre- and post-intervention periods. Our study was conducted during the COVID-19 pandemic. In Uganda, shelter-in-place went into effect in March 2020 and was associated with an immediate decline in TB case notifications.^28^ This was consistent with our data and may indicate a different population observed during the post-intervention period. However, if the population initiated on treatment after the shelter-in-place policies went into effect was sicker and at higher risk of poor outcomes due to selection bias, we would expect the impact of the intervention to be even greater than observed in our study. In the post-intervention period, there was a higher proportion of females as well as a higher proportion being treated at a health center as compared to hospital, which may have also affected outcomes.

In summary, there was high uptake of 99DOTS among adolescents in this study, and the use of 99DOTS was associated with improved treatment outcomes. TB disease and treatment are known to negatively impact domains of well-being crucial to a successful transition between childhood and adulthood including good health, connectedness to society, education, and agency.^5,29^ DATs are a person-centered alternative to DOT for TB treatment that may optimize adolescents’ good health by improving treatment outcomes. Furthermore, by giving adolescents the ability to choose the time and location for their medication dosing, DATs may decrease stigma, minimize interruptions in school, and improve agency. These technologies should be further tailored towards adolescents in the effort to combat the global TB epidemic.

## Data Availability

All data produced in the present study are available upon reasonable request to the authors.

## APPENDIX

## Supplemental Methods. Equation for interrupted time series regression model

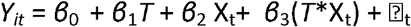

Where:

- *Y*_*it*_ is the aggregated outcome in a given health facility during each study month
- *T* is the time since the start of the study, X_t_ is the intervention period indicator variable coded 0 for pre-intervention and 1 for post-intervention, and T^*^X_t_ is an interaction term
- *β*_*0*_ is the intercept of the outcome for the pre-intervention period,
- *β*_*1*_ is the slope for the pre-intervention period,
- *β*_*2*_ is the level change following the intervention, with (*β*_*0*_ + *β*_*2*_) representing the intercept of the outcome for the post-intervention period,
- *β*_*3*_ is the difference in post-intervention and pre-intervention period slopes, with (*β*_*1*_ + *β*_*3*_) representing the slope of the post-intervention period

**Supplemental Figure 1.**
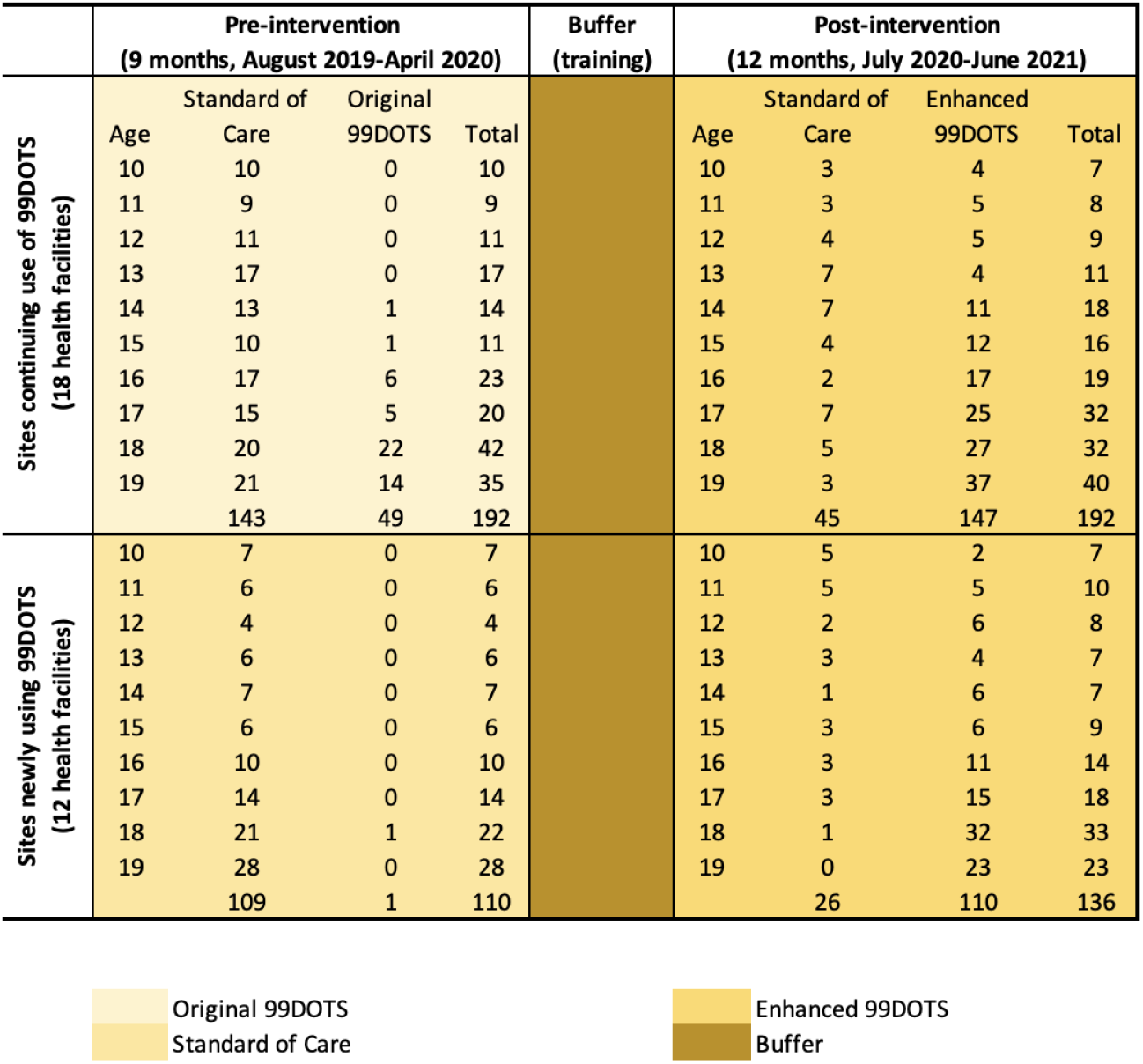
Interrupted time series study design and enrollment on 99DOTS.

**Supplemental Table 1.** Interrupted time series regression results from an unadjusted model in all adolescents.

|  | Change in level<br>(PR, 95% CI) | Change in slope<br>(PR, 95% CI) | Pre-intervention<br>slope (PR, 95% CI) | Post-intervention<br>slope (PR, 95% CI) |
| --- | --- | --- | --- | --- |
| Treated successfully |  |  |  |  |
| ITT | 1.16 (1.06-1.29)* | 1.01 (0.99-1.03) | 0.99 (0.97-1.00) | 1.00 (0.99-1.00) |
| PP | 1.16 (1.05-1.28)* | 1.01 (0.99-1.03) | 0.99 (0.97-1.00) | 1.00 (0.98-1.01) |
| Loss to follow-up |  |  |  |  |
| ITT | 0.92 (0.87-0.97)* | 1.00 (0.99-1.01) | 1.01 (1.00-1.01) | 1.01 (1.00-1.01) |
| PP | 0.93 (0.88-0.98)* | 1.00 (0.99-1.01) | 1.01 (1.00-1.01) | 1.00 (0.99-1.01) |

PR: proportion ratio; CI: confidence interval; ITT: intention-to-treat; PP: per protocol analysis Statistically significant results noted with \* (<0.05)

**Supplemental table 2.** Interrupted time series regression results from an unadjusted model in older adolescents (ages 15-19).

|  | Change in level<br>(PR, 95% CI) | Change in slope<br>(PR, 95% CI) | Pre-intervention<br>slope (PR, 95% CI) | Post-intervention<br>slope (PR, 95% CI) |
| --- | --- | --- | --- | --- |
| Treated successfully |  |  |  |  |
| ITT | 1.26 (1.11-1.43)* | 1.02 (1.00-1.04) | 0.98 (0.96-1.00)* | 0.99 (0.99-1.00) |
| PP | 1.25 (1.09-1.42)* | 1.02 (1.00-1.04) | 0.98 (0.96-1.00)* | 1.00 (0.99-1.01) |
| Loss to follow-up |  |  |  |  |
| ITT | 0.88 (0.81-0.94)* | 1.00 (0.99-1.01) | 1.01 (1.00-1.02) | 1.01 (1.00-1.01)* |
| PP | 0.88 (0.82-0.95)* | 0.99 (0.98-1.00) | 1.01 (1.00-1.02) | 1.00 (0.99-1.01) |

PR: proportion ratio; CI: confidence interval; ITT: intention-to-treat; PP: per protocol analysis Statistically significant results noted with \* (<0.05)

**Supplemental Figure 2.**
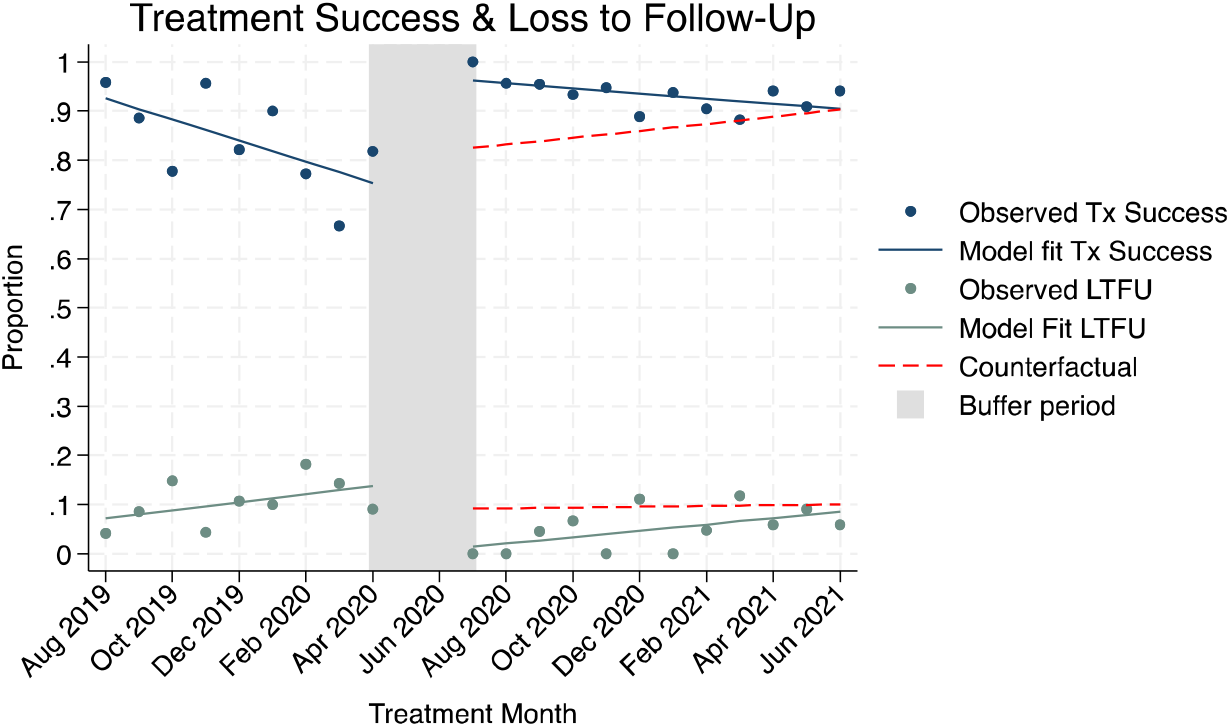
Treatment success and loss to follow-up for older adolescents (ages 15-19).

## REFERENCES

1. Snow KJ, Cruz AT, Seddon JA, et al. Adolescent tuberculosis. Lancet Child Adolesc Health. 2020;4(1):68–79. doi:10.1016/S2352-4642(19)30337-2

2. Gupta KB, Gupta R, Atreja A, Verma M, Vishvkarma S. Tuberculosis and nutrition. Lung India Off Organ Indian Chest Soc. 2009;26(1):9–16. doi:10.4103/0970-2113.45198

3. Kant S, Gupta H, Ahluwalia S. Significance of Nutrition in Pulmonary Tuberculosis. Crit Rev Food Sci Nutr. 2015;55(7):955–963. doi:10.1080/10408398.2012.679500

4. Christian P, Smith ER. Adolescent Undernutrition: Global Burden, Physiology, and Nutritional Risks. Ann Nutr Metab. 2018;72(4):316–328. doi:10.1159/000488865

5. Moscibrodzki P, Enane LA, Hoddinott G, et al. The Impact of Tuberculosis on the Well-Being of Adolescents and Young Adults. Pathogens. 2021;10(12):1591. doi:10.3390/pathogens10121591

6. Yellappa V, Lefèvre P, Battaglioli T, Narayanan D, Van der Stuyft P. Coping with tuberculosis and directly observed treatment: a qualitative study among patients from South India. BMC Health Serv Res. 2016;16:283. doi:10.1186/s12913-016-1545-9

7. Chiang SS, Senador L, Altamirano E, et al. Adolescent, caregiver and provider perspectives on tuberculosis treatment adherence: a qualitative study from Lima, Peru. BMJ Open. 2023;13(5):e069938. doi:10.1136/bmjopen-2022-069938

8. Zhang S, Li X, Zhang T, Fan Y, Li Y. The experiences of high school students with pulmonary tuberculosis in China: a qualitative study. BMC Infect Dis. 2016;16:758. doi:10.1186/s12879-016-2077-y

9. Enane LA, Lowenthal ED, Arscott-Mills T, et al. Loss to follow-up among adolescents with tuberculosis in Gaborone, Botswana. Int J Tuberc Lung Dis. 2016;20(10):1320–1325. doi:10.5588/ijtld.16.0060

10. Snow KJ, Sismanidis C, Denholm J, Sawyer SM, Graham SM. The incidence of tuberculosis among adolescents and young adults: a global estimate. Eur Respir J. 2018;51(2):1702352. doi:10.1183/13993003.02352-2017

11. Wobudeya E, Jaganath D, Sekadde MP, Nsangi B, Haq H, Cattamanchi A. Outcomes of empiric treatment for pediatric tuberculosis, Kampala, Uganda, 2010–2015. BMC Public Health. 2019;19:446. doi:10.1186/s12889-019-6821-2

12. Osman M, du Preez K, Seddon JA, et al. Mortality in South African Children and Adolescents Routinely Treated for Tuberculosis. Pediatrics. 2021;147(4):e2020032490. doi:10.1542/peds.2020-032490

13. Leddy AM, Jaganath D, Triasih R, et al. Social Determinants of Adherence to Treatment for Tuberculosis Infection and Disease Among Children, Adolescents, and Young Adults: A Narrative Review. J Pediatr Infect Dis Soc. 2022;11(Supplement_3):S79-S84. doi:10.1093/jpids/piac058

14. Chiang SS, Waterous PM, Atieno VF, et al. Caring for Adolescents and Young Adults with Tuberculosis or at Risk of Tuberculosis: Consensus Statement from an International Expert Panel. J Adolesc Health Off Publ Soc Adolesc Med. 2023;72(3):323–331. doi:10.1016/j.jadohealth.2022.10.036

15. Swahn MH, Braunstein S, Kasirye R. Demographic and Psychosocial Characteristics of Mobile Phone Ownership and Usage among Youth Living in the Slums of Kampala, Uganda. West J Emerg Med. 2014;15(5):600–603. doi:10.5811/westjem.2014.4.20879

16. Cattamanchi A, Crowder R, Kityamuwesi A, et al. Digital adherence technology for tuberculosis treatment supervision: A stepped-wedge cluster-randomized trial in Uganda. PLoS Med. 2021;18(5):e1003628. doi:10.1371/journal.pmed.1003628

17. Chen AZ, Kumar R, Baria RK, Shridhar PK, Subbaraman R, Thies W. Impact of the 99DOTS digital adherence technology on tuberculosis treatment outcomes in North India: a pre-post study. BMC Infect Dis. 2023;23:504. doi:10.1186/s12879-023-08418-2

18. Cross A, Gupta N, Liu B, et al. 99DOTS: a low-cost approach to monitoring and improving medication adherence. In: ; 2019. Accessed August 18, 2023. https://www.researchgate.net/publication/330272562_99DOTS_a_low-cost_approach_to_monitoring_and_improving_medication_adherence

19. Yoeli E, Rathauser J, Bhanot SP, et al. Digital Health Support in Treatment for Tuberculosis. N Engl J Med. 2019;381(10):986–987. doi:10.1056/NEJMc1806550

20. Crowder R, Nakasendwa S, Kityamuwesi A, et al. Implementation of enhanced 99DOTS for TB treatment supervision in Uganda: An interrupted time series analysis. Published online January 23, 2024:2024.01.22.24300949. doi:10.1101/2024.01.22.24300949

21. Roadmap towards Ending TB in Children and Adolescents, Third Edition. World Health Organization; 2023.

22. Crowder R, Kityamuwesi A, Kiwanuka N, et al. Study protocol and implementation details for a pragmatic, stepped-wedge cluster randomised trial of a digital adherence technology to facilitate tuberculosis treatment completion. BMJ Open. 2020;10(11):e039895. doi:10.1136/bmjopen-2020-039895

23. Berger CA, Kityamuwesi A, Crowder R, et al. Variation in tuberculosis treatment outcomes and treatment supervision practices in Uganda. J Clin Tuberc Mycobact Dis. 2020;21:100184. doi:10.1016/j.jctube.2020.100184

24. Harris PA, Taylor R, Thielke R, Payne J, Gonzalez N, Conde JG. Research Electronic Data Capture (REDCap) - A metadata-driven methodology and workflow process for providing translational research informatics support. J Biomed Inform. 2009;42(2):377–381. doi:10.1016/j.jbi.2008.08.010

25. Linden A. Conducting Interrupted Time-series Analysis for Single- and Multiple-group Comparisons. Stata J. 2015;15(2):480–500. doi:10.1177/1536867X1501500208

26. Ridho A, Alfian SD, van Boven JFM, et al. Digital Health Technologies to Improve Medication Adherence and Treatment Outcomes in Patients With Tuberculosis: Systematic Review of Randomized Controlled Trials. J Med Internet Res. 2022;24(2):e33062. doi:10.2196/33062

27. Kinghorn A, Shanaube K, Toska E, Cluver L, Bekker LG. Defining adolescence: priorities from a global health perspective. Lancet Child Adolesc Health. 2018;2(5):e10. doi:10.1016/S2352-4642(18)30096-8

28. Kadota JL, Reza TF, Nalugwa T, et al. Impact of shelter-in-place on TB case notifications and mortality during the COVID-19 pandemic. Int J Tuberc Lung Dis Off J Int Union Tuberc Lung Dis. 2020;24(11):1212–1214. doi:10.5588/ijtld.20.0626

29. Ross DA, Hinton R, Melles-Brewer M, et al. Adolescent Well-Being: A Definition and Conceptual Framework. J Adolesc Health. 2020;67(4):472–476. doi:10.1016/j.jadohealth.2020.06.042

